# A Prospective Pilot Clinical Study reveals a promising non-toxic anti-biofilm activity of Gentamicin-EDTA-Na₂ Central Venous Catheter Lock Solution

**DOI:** 10.1101/2025.03.21.25324295

**Authors:** David Lebeaux, Bérénice Souhail, Tan-Phuc Bui Van, Lénaig Le Fouler, Matthieu Lafaurie, Raphael Lepeule, Etienne Canoui, Victoire de Lastours, Antoine Froissart, Damien Blez, Christophe Beloin, Jean-Marc Ghigo, Fabrice Pirot, Carole Dhelens, Sandrine Fernandes-Pellerin

## Abstract

**Objectives:** The treatment of long-term intravenous catheter-related bloodstream infections (LTIVC-related BSI) often requires catheter removal or a conservative treatment using intra-catheter locks, with a 50-60% success rate. We previously demonstrated the synergistic effect of a combination of gentamicin and EDTA-Na_2_ against bacterial biofilms. We performed a phase 1/2 clinical trial to assess the tolerance and efficacy of the genta-EDTA-Na_2_ locks for the conservative treatment of LTIVC-related BSI.

**Methods:** Prospective study including adult patients with a monomicrobial uncomplicated LTIVC-related BSI caused by a gentamicin-susceptible coagulase-negative staphylococci, *Enterobacterales* or *Pseudomonas aeruginosa*. Primary objective: assess the safety and efficacy at Day 40 (D40) of genta-EDTA-Na_2_ locks by evaluating the frequency of clinical and microbiological cure 30 days after the end of treatment (D40).

**Results:** Eight patients were included. A complete follow-up was obtained for 7 patients, 6 of which met cure criteria. The single patient whose follow-up was incomplete met all criteria for cure at D23. A single microbiological failure occurred (relapse of *P. aeruginosa* LTIVC-related BSI). Two patients experienced at least one serious adverse event; none were attributed to the genta-EDTA-Na_2_ locks.

**Conclusion:** Genta-EDTA-Na_2_ used as intra-catheter locks is a promising anti-biofilm candidate to be studied in a randomized controlled trial.

## INTRODUCTION

Long-term intravenous catheters (LTIVC) are crucial for millions of patients dependent upon hemodialysis, parenteral nutrition or antineoplastic chemotherapy, but their use can be complicated by catheter-related bloodstream infections (LTIVC-related BSI) (1). Treatment of LTIVC-related BSI relies on catheter removal or, alternatively, a conservative strategy combining systemic antibiotics and intra-catheter locks, using a small volume (2-3 mL) of highly-concentrated antibiotic (100 to 1000 times the minimal inhibitory concentration) to eradicate bacterial biofilms colonizing the catheter (1). However, the success rate of the currently used antibiotic locks is limited to 50-60% (2, 3).

We demonstrated that the combination of ethylenediaminetetraacetic acid disodium salt (EDTA-Na_2_) and the aminoglycoside antibiotic gentamicin eradicated *in vivo* biofilms in a rat model of LTIVC-related infections and was synergistic against a broad-range of bacterial isolates responsible for LTIVC-related infections (4, 5). Subsequently, a study using commercial gentamicin sulfate and EDTA-Na_2_ the long-term stability of genta-EDTA-Na_2_ lock solutions for 12 months (6).

Here, we performed a pilot phase 1/2 clinical trial to assess the tolerance and efficacy of genta-EDTA-Na_2_ used as an intra-catheter lock for the curative and conservative treatment of LTIVC-related BSI.

## MATERIAL AND METHODS

### Study setting and inclusion criteria

We performed a prospective interventional study (CATH-GE) between April 2021 and November 2023 in five tertiary hospitals of the Ile-de-France region, France (**Supplementary Table 1**) (ClinicalTrials.gov Identifier: NCT04789928). Adult patients were included if they had an uncomplicated LTIVC-related BSI eligible for a conservative treatment (**Supplementary Methods)**.

### Objectives and endpoints

The primary objective was to assess the efficacy and safety at Day 40 (D40) of genta-EDTA-Na_2_ used as an intra-catheter lock for the conservative treatment of uncomplicated LTIVC-related BSI. The efficacy was assessed by the frequency of clinical and microbiological cure 30 days after the end of treatment (D40) (**Supplementary Methods**). The safety of genta-EDTA-Na_2_ locks was assessed by the frequency of any suspected related adverse event (**Supplementary Methods)**.

### Study design and follow-up

This was a pilot phase, non-comparative, non-randomized, multicentric study. We initially planned to include 35 patients as we expected to describe a 90% clinical and microbiological cure rate at D40 with a 10% precision and an alpha-risk of 0.05. However, due to major inclusion limitations, we stopped inclusion in November 2023.

The study design included: *i)* a screening and inclusion visit that occurred within the 72 hours following the day the first blood culture drawn from the LTIVC was positive; *ii)* 7 to 10 days genta-EDTA-Na_2_ lock treatment (D6-D9) depending on the prescription of a previous treatment lock to the patient (see **Supplementary Figure 1**), *iii)* a daily clinical and biological (see below) follow-up during the genta-EDTA-Na_2_ lock treatment; *iv)* an efficacy and safety follow-up visit at D40 (+/− 7 days); *v)* an end of study visit at D100 (+/− 7 days).

Day 0 (D0) was defined by the initiation of genta-EDTA-Na_2_ lock therapy. On D1, D2 and D6/9, blood samples (blood ionized calcium level, gentamicin concentration and serum creatinine) were performed from a peripheral vein to avoid contamination by previous lock.

Control blood cultures were sampled from a peripheral vein and from the LTIVC on D3/5, D6/9 and on D40. Venous ultrasound was performed between D2 and D5 and transthoracic echocardiography was performed between D2 and D7.

Microbiological relapse was defined as a BSI with the same microbial pathogen, *i.e.* same species and same antibiotic susceptibility pattern.

### Production of genta-EDTA-Na_2_ vials and description of the use of the intra-catheter lock solution

The production and labeling of the genta-EDTA-Na_2_ solution were carried out by the Edouard Herriot Hospital Pharmacy, on the FRIPHARM® platform, Hospices Civils de Lyon, as previously described (6) (**Supplementary Methods**). Intra-catheter locks were performed *as per* routine care (**Supplementary Methods**): briefly, the catheter dead space was measured for the first use. Each lock was performed with dead space volume + 0.5 mL. Each lock was maintained for 24 hrs. Twenty-four hours later, previous lock (at least 3 mL) was aspirated and then replaced by a new genta-EDTA-Na_2_ lock.

## Collected Data: see supplementary Method

### Statistical Analysis

The proportion of clinical and microbial cure at D40 and D100 and the proportion of suspected related adverse events were assessed. Considering the low number of included patients, no statistical analysis was performed.

### Ethics

The trial was conducted in compliance with the principles of Good Clinical Practice and the Declaration of Helsinki for biomedical research involving human beings. It was approved by the French National Ethics Committee (Comité de Protection des Personnes, CPP Île de France 11) and authorized by the Agence nationale de sécurité du médicament et des produits de santé. A written informed consent was obtained from all patients enrolled in the trial. The trial was registered at Clinical-Trials.gov (NCT04789928).

## RESULTS

### Characteristics of Patients

Eight patients were included with a median age of 66 [36-83] years (**Table 1**). All patients had TIVAP, mostly for antineoplastic chemotherapy (7/8). Three infections were caused by *P. aeruginosa*, 3 by coagulase negative staphylococci and 2 by *Enterobacterales*.

**Table 1.** Clinical characteristics and outcome of eight patients receiving genta-EDTA-Na_2_ as intra-catheter locks.

| Patient number | Gender | Age range, year | Reason for LTIVC implantation | Microbial pathogen | Initial eGFR | Total duration of lock (days) | Total duration of genta-EDTA-Na<sup>2</sup> lock (days) | Clinical and microbiological cure at D40 | Clinical and microbiological cure at D100 |
| --- | --- | --- | --- | --- | --- | --- | --- | --- | --- |
| LCK-1-001 | Male | 36-40 | Cancer | *P. aeruginosa* | 73 | 10 | 10 | NA* | NA* |
| LCK-2-001 | Female | 81-85 | Cancer | *E. coli* | 33 | 9 | 9 | yes | yes |
| LCK-2-002 | Female | 71-75 | Parenteral nutrition | *E. cloacae* | 98 | 9 | 9 | yes | yes |
| LCK-3-001 | Female | 76-80 | Cancer | *S. epidermidis* | 83 | 10 | 8 | yes | yes |
| LCK-3-002 | Female | 61-65 | Cancer | *S. hominis* | 80 | 10 | 8 | yes | yes |
| LCK-3-003 | Male | 46-50 | Cancer | *P. aeruginosa* | 130 | 10 | 7 | no** | no** |
| LCK-3-004 | Female | 76-80 | Cancer | *P. aeruginosa* | 66 | 10 | 7 | yes | yes |
| LCK-3-005 | Male | 61-65 | Cancer | *S. haemolyticus* | 128 | 10 | 9 | yes | yes |

**NOTE:** eGFR: estimated Glomerular Filtration Rate, in mL/min/1.73m<sup>2</sup> ; NA: not available.
\*Patient LCK-1-001 had negative paired blood cultures performed at D3, D7 and D14. His last clinical follow-up was on D23 when he met all criteria for clinical and microbiological cure. He was then transferred to his native country for palliative care.
\*\*Patient LCK-3-003 experienced a relapse of his LTIVC-related BSI with the same bacterial species (*P. aeruginosa*).

### Efficacy of genta-EDTA-Na_2_ intra-catheter locks

A complete follow-up was obtained at day 40 for seven patients, six of which met clinical and microbiological cure criteria with negative blood cultures at D40 (+/− 7 days) (**Table 1**). The single patient whose follow-up was incomplete (LCK-1-001) had negative paired blood cultures performed at D3, D7 and D14 and his last clinical follow-up was on D23 when he met all criteria for clinical and microbiological cure. All patients meeting clinical and microbiological cure criteria at day 40 also met these criteria at day 100. A single microbiological failure occurred (LCK-3-003) who experienced a relapse of his LTIVC-related BSI (*P. aeruginosa*) with no local nor systemic complication.

### Tolerance of genta-EDTA-Na_2_ intra-catheter locks

Between D0 and D40, 2/8 patients experienced at least one serious adverse event (**Supplementary Table 2**). None were attributed to the genta-EDTA-Na_2_ intra-catheter lock. Follow-up blood ionized calcium and serum creatinine levels are depicted in **Figure 1A** and **1B**. All gentamicin dosages were negatives.

**Figure 1.**
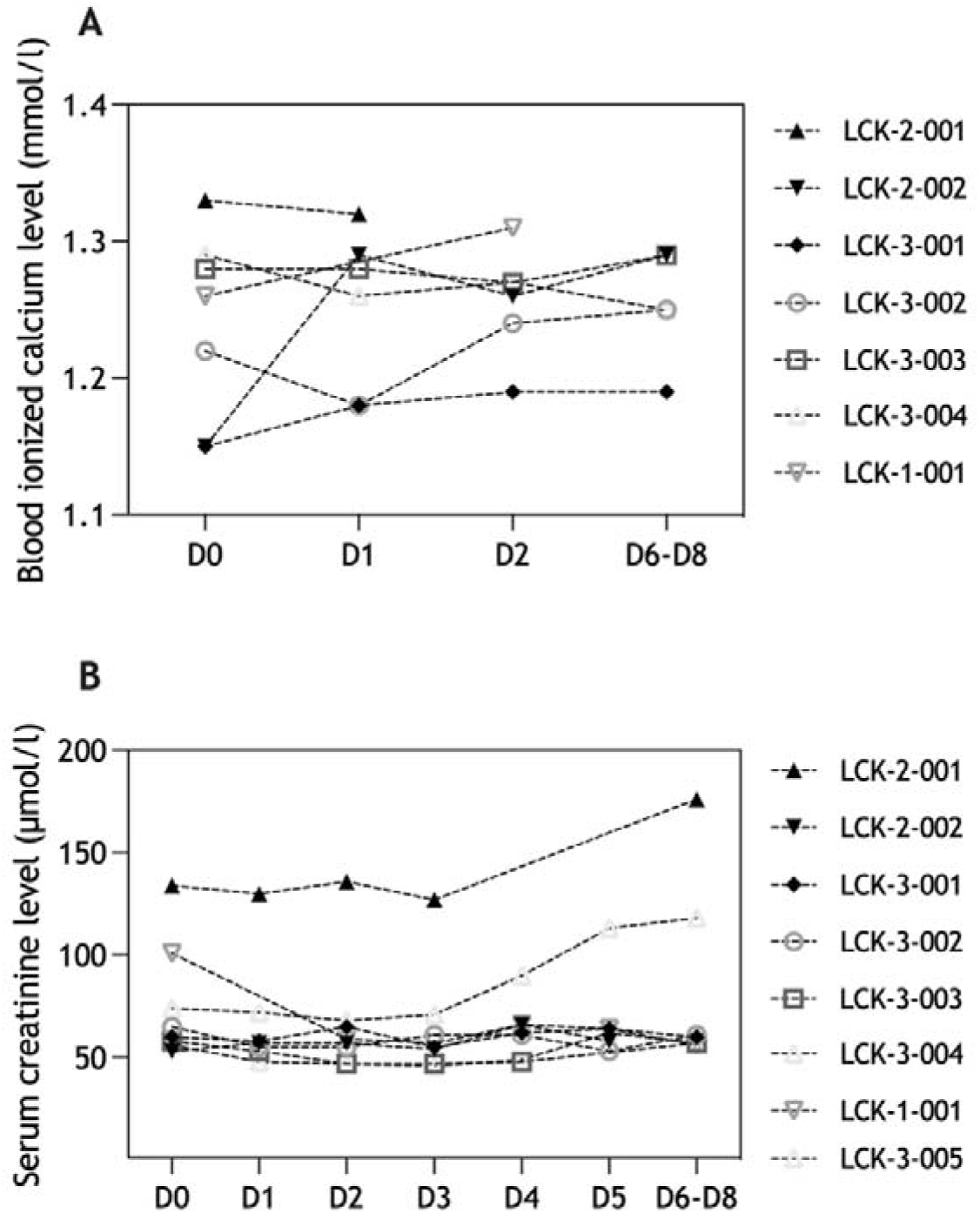
Evolution of blood ionized calcium (mmol/l) and serum creatinine (µmol/l) levels among eight patients receiving a genta-EDTA-Na_2_ intra-catheter lock. Patient LCK-1-001 never had a blood ionized calcium level measurement. His blood calcium levels remained stable along the treatment duration. Patient LCK-2-001 experienced an increase of her creatinine level after introduction of cotrimoxazole. Patient LCK-3-004 experienced an acute functional kidney failure secondary to cancer-induced diarrhea.

## DISCUSSION

In this pilot phase 1/2 clinical trial using genta-EDTA-Na_2_ as an intra-catheter lock, we observed no treatment-related adverse event, a favorable tolerance profile and a high efficacy for the conservative treatment of LTICV-related BSI (7 treatment successes/8 treated patients).

Recent data reported success rates following conservative treatment comprised between 50 and 60 % (2, 3). Our pilot study showed that 7 out of 8 patients experienced clinical and microbiological cure using genta-EDTA-Na_2_ as an intra-catheter lock, one having a limited follow-up of 23 days. In this context, the main limitation of our study is the small sample size, precluding to draw any definite conclusion regarding the efficacy of genta-EDTA-Na_2_ used as intra-catheter lock.

Regarding the safety profile of genta-EDTA-Na_2_ locks, we observed no adverse event related to the experimental treatment. No patient experienced a significant reduction of ionized calcium levels, and all gentamicin dosages were negatives. Two patients experienced an increase in their serum creatinine levels, none being related to the genta-EDTA-Na_2_ lock. Besides, the tolerance profile of EDTA used as intra-catheter lock was favourable among 190 patients receiving EDTA-minocycline as preventive lock in previously published studies (7-10).

In conclusion, the use genta-EDTA-Na_2_ as intra-catheter locks for the conservative treatment of LTIVC-related BSI appears to be a promising candidate to be studied in a randomized controlled trial.

## ACKNOWLEGEMENTS

The authors would like to thank Cécile Artaud, Simon Amador Paz and Charles-Henry Bourlier for their help in preparing the study protocol.

## Funding

The CATH-GE study has been funded as follows: Fonds de Dotation Patrick de Brou de Lauriere; Sauver la vie Grant (Paris Descartes); A.R.C.A.D (Aide à la Recherche en CAncérologie Digestive); Ligue contre le cancer; mécénat Air France, ANR-10-LABX-62-IBEID, Agence Nationale de la Recherche, Laboratoire d’Excellence “Integrative Biology of Emerging Infectious Diseases”. We also would like to thank all our friends, family and colleagues who supported this project through a Thellie Crowdfunding.

## Conflict of interest

The authors declare that they have no conflict of interest to declare

## Data Availability

All data produced in the present study are available upon reasonable request to the authors

## SUPPLEMENTARY MATERIALS

### Supplementary Methods

#### Comprehensive list of inclusion and non-inclusion criteria Inclusion criteria

- Adult patient (≥18 year-old);
- LTIVC in place (TIVAP or single-line tunnelled catheter);
- LTIVC is functional (it is possible to inject an infusate, but also to draw blood from the catheter);
- LTIVC-related bloodstream infection defined by a positive qualitative paired blood culture with a differential time to positivity ≥ 2 hours (meaning that the culture of the blood drawn from the catheter is positive at least 2 hours before the culture of the blood drawn from a peripheral vein);
- Mono microbial infection caused by coagulase-negative staphylococci, Enterobacteriaceae or Pseudomonas aeruginosa;
- Bacterial strain is susceptible to gentamicin;
- Life expectancy ≥ 3 months;
- Physician in charge of the patient agrees to perform a conservative treatment;
- estimated Glomerular Filtration Rate (eGFR) ≥ 30 mL/min/1.73m^2^;
- Patient’s informed and written consent is collected.
- For women of reproductive age: available beta-HCG dosage (with negative result) < 72h.

Non-inclusion criteria

- Presence of any systemic complication (sepsis or septic shock), or local complications (tunnel or port-pocket infection, thrombophlebitis, endocarditis, bone and joint infections related to the LTIVC-related BSI);
- Allergy to aminoglycosides;
- PICC-line or hemodialysis tunnelled catheter;
- LTIVC removal is planned within the following 3 months or LTIVC is not required for the management of the patient’s underlying medical condition anymore;
- Diagnosis of LTIVC-related bloodstream infection has been made more than 3 days ago (e.g. >72 hours between the day the first blood culture drawn from the LTIVC is positive and the screening visit);
- Systemic treatment of LTIVC-related bloodstream infection includes aminoglycosides (defined as a recent (<36 hours) or ongoing systemic injection of aminoglycosides)
- Low blood ionized calcium level (<1,15 mmol/L) before injecting the first dose of genta-EDTA-Na_2_ lock;
- Presence of prosthetic heart valve, pacemaker or implantable defibrillator;
- The LTIVC has been inserted less than 14 days ago;
- Available Count blood cells < 72h with severe neutropenia (<500 polymorphonuclear cells/mm);
- Subject with infection caused by Staphylococcus aureus or Candida spp.;
- The patient is not expected to remain in hospital for at least 7 days after inclusion
- The administration of the lock according to the protocol (24 hours/day for 48 hours and then at least 12 hours/24 hours for 5 to 8 days) is not possible.
- Decision by the referring physician to prescribe a preventive lock following curative Gentamicin-EDTA treatment (secondary prevention), between the end of treatment (D6/D9) and the D40 follow-up visit.
- Previous inclusion in a study or another therapeutic protocol requiring continuous use of the catheter
- Inability to perform a sampling of peripheral venous blood
- Pregnant and breastfeeding women,
- Protected adult subject

### Exclusion criteria

A patient should be excluded if he presents any of the following criteria:

– Presence of local or systemic complication seen on venous ultrasound performed between D2 and D5 or transthoracic echocardiography performed between Day 2 and Day 7 (D2-D7);
– GFR < 30 ml/min (between D0 and D6/9). The subject will not be excluded if another medical condition is identified as responsible for this decrease.

#### Comprehensive definition of clinical and microbiological cure

The **efficacy** was assessed by the frequency of **clinical and microbiological cure** after 30 days of follow-up after the end of treatment (D40).

**Clinical and microbiological cure was defined by** the completion of all the following criteria:

- Resolution of clinical signs at the end of treatment (D6 if the patient already received 72h of a previous active lock before inclusion or D9 if he received a complete 10-day course of genta-EDTA-Na locks) and 30 days after completion of therapy (D40) (see **Supplementary Figure 1** for the definition of days after inclusion)
- Negative blood cultures during treatment:
  ○ drawn from a peripheral vein (D3-D5);
  ○ drawn from a peripheral vein and from LTIVC (EOT = D6-D9);
- Negative blood cultures drawn 30 days after the end of treatment (D40). If initial infection was caused by coagulase-negative staphylococci, two sets of blood cultures sampled at 2 different moments were required to confirm failure of conservative treatment. Indeed, a single positive blood culture sampled on LTIVC or a peripheral vein may be related to a microbial contamination of blood culture bottles during the sampling procedure;
- Absence of microbiological relapse (BSI with the same microbial pathogen, i.e. same species and same antibiotic susceptibility pattern) between D6-D9 and D40.
- No occurrence of endocarditis, thrombophlebitis or osteomyelitis caused by the LTIVC-related BSI during follow-up;
- Absence of catheter dysfunction between D0 and D6-D9

The occurrence of a new LTIVC-related infection caused by another microbial pathogen (different species or different antibiotic susceptibility pattern in case of identical species) was not considered as a microbiological failure. The occurrence of a mechanical dysfunction (i.e. unrelated to the infection) after D6-D9 was not considered as a failure (see below).

#### Comprehensive definitions of adverse events

##### □ Adverse event

Any untoward medical occurrence, unintended disease or injury or any untoward clinical signs (including an abnormal laboratory finding) in subjects, users or other persons whether or not related to the investigational treatment.

– Note 1: This includes events related to the investigational treatment
– Note 2: This includes events related to the procedures involved (any procedure in the clinical investigation plan).

All adverse events experienced during the study, which are observed by the investigator or reported by the patient, will be recorded in the section of the case report form provided for this purpose.

For all events the scoring will be as follows according to the CTCAE:

1 = mild; 2 = moderate; 3 = severe; 4 = life-threatening; 5= death

##### □ Adverse reaction

Any adverse event for which a causality link may be possible with the study treatment, or study-specific procedures, regardless of its significance (doubtful, plausible, possible, sure).

The procedures performed as part of the medical care are not considered as required by the protocol because they would have been carried out whether the patient participated in the study or not. In this particular case, the adverse events linked to these procedures will not be considered as adverse reactions.

##### □ Serious adverse event

An adverse event/reaction must be considered as serious when it:

- results in death,
- is life-threatening,
- requires hospitalisation or prolongation of existing prolongation,
- results in persistent or significant disability or incapacity,
- is a congenital anomaly or birth defect,
- is medically important (Some medical events may jeopardise the subject or may require an intervention to prevent one of the above characteristics/consequences.).

The following events will not be considered as serious:

- A planned hospitalization for pre-existing condition, or a study-specific procedure described in the protocol, without a serious deterioration in health
- Hospitalisation for a duration less than 24 h (i.e. not requiring a night in the hospital), unless it could be considered as an important medical event,
- Hospitalisations linked to the studied pathology, including infection relapse or failure to reach microbial eradication
- Hospitalisations which have been planned before the subject’s participation in the protocol,
- Hospitalisation for the treatment of a concomitant pathology diagnosed before the subject’s participation in the protocol, except if the treatment intensity or frequency is increased during the subject’s participation in the protocol,
- Hospitalisation for routine clinical procedures as well as on social grounds.

##### □ Expected adverse reaction

An expected adverse reaction is a reaction:

- Already mentioned in the most recent versions of the investigator’s brochure or in the Summary of Product Characteristics (SPC) for drugs that already have a marketing authorisation or already identified during the practice of the procedures planned as part of this project.
- Foreseeable and linked to examinations and samplings required by the protocol (e.g. for the taking of the blood sample: pain, bruising, fainting spell).

##### □ Expected Serious Adverse Reactions (ESAR)

ESAR will be defined as follow:

– For examinations and samplings required by the protocol: Consequences of vasovagal response due to repeated blood sampling.
– Serious adverse reactions mentioned in the reference safety information (RSI) of gentamicin: gentamicin’s SmPC
– Serious adverse reactions mentioned in the RSI of EDTA Na_2:_ EDTA Na_2_’s SmPC

#### Comprehensive description of production of Genta-EDTA-Na^2^ vials

The production of genta-EDTA-Na solution conditioned in sealed glass containers (cf. 3.2.1 European Pharmacopoeia monograph, Ed. 9.4) was performed at the Edouard Herriot Hospital Pharmacy, on FRIPHARM platform, Hospices Civils de Lyon, which is authorized for the production and control of experimental pharmaceutical batches, according to GMP (Bonnes Pratiques de Préparation, ANSM, 2023). Genta-EDTA-Na_2_ lock solutions were obtained by compounding commercial gentamicin sulfate (Gentamicin PANPHARM® 80 mg, solution for injection) and EDTA-Na_2_ in aseptic conditions in sterile and non-pyrogenic amber glass type I pharmaceutical vials followed by sterilizing filtration and terminal sterilization by autoclaving. Particulate contamination by sub-visible particles was assessed as described in 2.9.19 European Pharmacopoeia monograph, Ed. 9.4. The sterility of genta-EDTA-Na solution was controlled as described in 2.6.1 European Pharmacopoeia monograph (Ed. 9.4). The number of units controlled for sterility was determined in function of the number of items in the batch as described below:

**Table 2.**
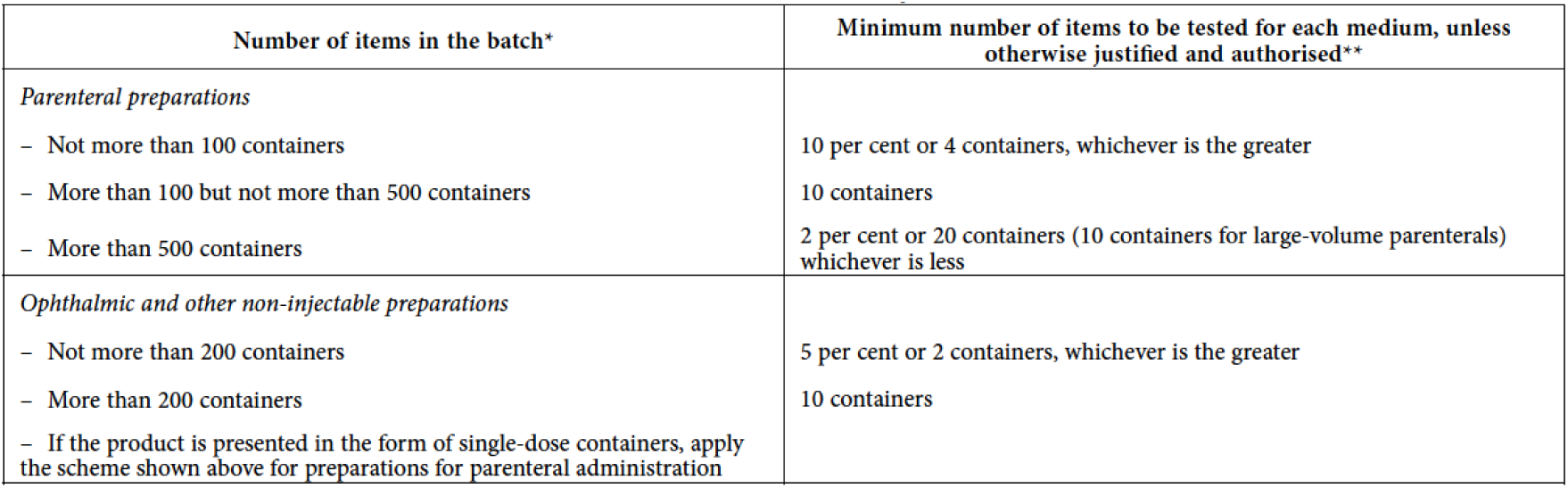
6.1.-3 – Minimum number of items to be tested.

The apyrogenicity of lock solution was assessed according to 5.1.10 European Pharmacopoeia monograph, Ed. 9.4. The endotoxin limit for active substances administered parenterally, defined on the basis of dose, is equal to: K/M

K = threshold pyrogenic dose of endotoxin per kilogram of body mass (Intravenous route: K = 5.0 IU of endotoxin per kilogram of body mass)

M = maximum recommended bolus dose of product per kilogram of body mass.

When the product is to be injected at frequent intervals or infused continuously, M is the maximum total dose administered per hour.

The endotoxin limit for lock solution was 0,5 IU/mL.

- The uniformity of content of single-dose preparation in 10 vials was determined according to 2.9.6 European Pharmacopoeia monograph, Ed. 9.4.

#### Comprehensive description of vial packaging

The vials were packaged in boxes of 15 and labeled in accordance with the French Decree of May 24, 2006, specifying the labeling requirements for investigational medicinal products. They were stored at room temperature with a maximum shelf life of 12 months and distributed by FRIPHARM® to investigational centers during the inclusion period

#### Description of the use of the intra-catheter lock solution

Patients included in this study received daily injection of genta-EDTA-Na lock associated with systemic antibiotics as follow:

- For the first use, nurse measured the catheter dead space by injecting 5 mL of sterile saline in the catheter and then drew blood with a 10 mL sterile syringe until recovering all the sterile saline solution. Note: Dead space volume (Vd) corresponds to the volume of sterile saline recovered in the syringe before collecting blood in the syringe. This volume was noted in the medical chart of the patient and collected in the eCRF.
- Further locks were performed with the following volume: Vd + 0.5 mL. In case of TIVAP, the complete procedure was performed with the Huber needle connected to a gripper.
- Then, the genta-EDTA-Na^2^ (Vd + 0.5 mL) lock was injected and maintained for 24hrs.
- Twenty-four hours later, previous lock (at least 3 mL) was aspirated and then replaced by a new genta-EDTA-Na lock.
- After the first 48 hours, if the patient reached apyrexia, locks could be changed each 12 hours, allowing antibiotic (or any other treatment) IV injections twice a day, through the LTIVC.

Blood cultures were scheduled at D3-D5 and at EOT (D6-D9); however additional blood cultures could be drawn on LTIVC and on peripheral vein if fever persisted after >48h of treatment. Blood collection was done as follows:

- On LTIVC: 5ml drawn and discarded in order to remove any trace of antibiotic from previous lock administration if any, then 5 ml on one aerobic bottle and 5 ml on one anaerobic bottle;
- From peripheral veins: 5 ml on one aerobic bottle and 5 ml on one anaerobic bottle

### Data collection tool and collected data

Study data were collected and managed using REDCap electronic data capture tools hosted at Institut Pasteur. REDCap (Research Electronic Data Capture) is a secure, web-based software platform designed to support data capture for research studies, providing 1) an intuitive interface for validated data capture; 2) audit trails for tracking data manipulation and export procedures; 3) automated export procedures for seamless data downloads to common statistical packages; and 4) procedures for data integration and interoperability with external sources.

Study data were collected and managed using REDCap electronic data capture tools hosted at Institut Pasteur. We collected the following data: demographic data; patient general condition (Charlson comorbidity and Karnofsky indexes); LTIVC (type, date of insertion, site, main reason for insertion: antineoplastic chemotherapy, parenteral nutrition, hemodialysis); underlying disease requiring the use of LTIVC; current use of parenteral nutrition.

We also recorded data regarding the current infection (date of the first symptoms; clinical signs and symptoms at diagnosis, biochemical and haematological characteristics at D0: C-reactive protein, leucocytes, neutrophils, creatinine level and clearance); radiological findings (venous ultrasound and transthoracic echocardiography).

Lastly, we collected data regarding the systemic (molecule, dose and duration) and local treatment (date of lock initiation, including previous lock, before inclusion, molecule used as lock before the initiation of genta-EDTA-Na_2_ lock) as well as clinical (cure, death, relapse as defined in **Supplementary methods** at 3 months after the end of treatment) and microbiological (results of follow-up blood cultures) outcome. Microbiological relapse was defined as a BSI with the same microbial pathogen, i.e. same species and same antibiotic susceptibility pattern.

## SUPPLEMENTARY TABLES

**Supplementary Table 1.** Comprehensive list of study centres.

|  | Principal investigator | Clinical<br>Microbiologist | Pharmacist |
| --- | --- | --- | --- |
| Hôpital Lariboisière, Paris<br>France | David Lebeaux | Ghalia Sbaa | Aude Jacob<br>Anas Gahbiche |
| Hôpital Européen Georges<br>Pompidou, Paris, France<br>Unité Mobile d'Infectiologie,<br>Service de Microbiologie | Damien Blez | Anne-Laure Bidaud | Brigitte Sabatier |
| Hôpital Saint-Louis, Paris,<br>France<br>Service des maladies<br>infectieuses | Matthieu Lafaurie | Béatrice Bercot | Sophie Touratier |
| Hôpital Henri Mondor,<br>Créteil, France<br>Unité Mobile d'Infectiologie,<br>Service de Microbiologie | Raphael Lepeule and<br>Berenice Souhail | Paul-Louis Woerther | Muriel Paul |
| Hôpital Cochin, Paris, France<br>Services des maladies<br>infectieuses et tropicales | Etienne Canoui | Hélène Poupet | Corinne Guérin |
| Hôpital Beaujon, Clichy,<br>France<br>Médecine Interne | Andréa De la Selle | Véronique Leflon | Zeina Louis |
| Centre Hospitalier Inter-Communal de Créteil, France<br>Service de médecine Interne | Antoine Froissart | Emmanuelle Varon | Stéphanie<br>Poullain |

**Supplementary Table 2.** Comprehensive list of adverse events reported among eight patients receiving genta-EDTA-Na_2_ as intra-catheter locks.

| Patient number | Any adverse event | Any serious adverse event | Description of the serious adverse event | Any suspected related adverse event |
| --- | --- | --- | --- | --- |
| LCK-1-001 | no | - |  | no |
| LCK-2-001 | yes | no |  | no |
| LCK-2-002 | no | - |  | no |
| LCK-3-001 | no | - |  | no |
| LCK-3-002 | yes | yes | Pulmonary embolism | no |
| LCK-3-003 | yes | no |  | no |
| LCK-3-004 | yes | yes | Acute functional kidney failure | no |
| LCK-3-005 | yes | no |  | no |

## SUPPLEMENTARY FIGURES

**Supplementary Figure 1.**
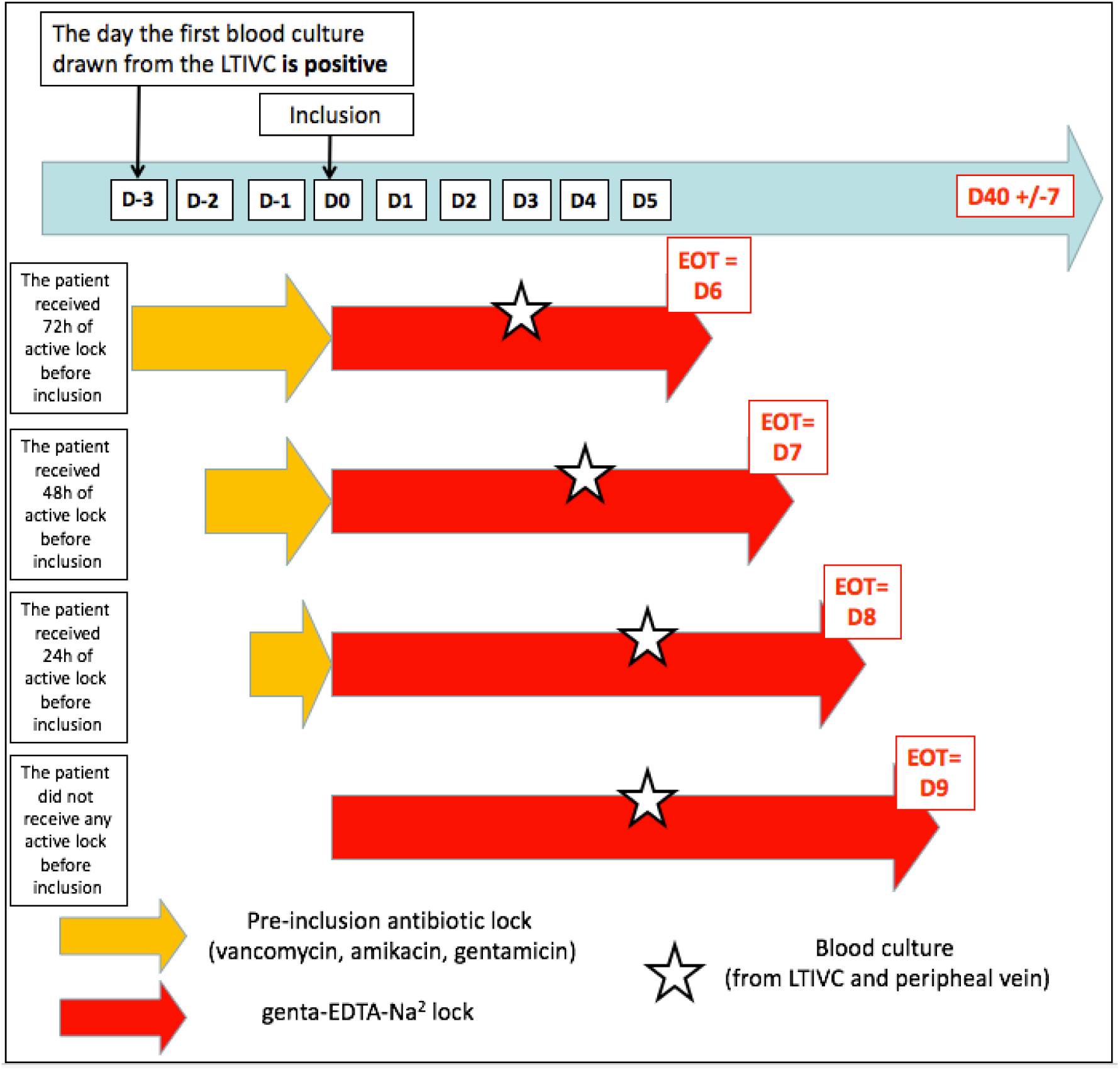
Definition of genta-EDTA-Na treatment duration after inclusion, depending on the timing between the initiation of any previous local treatment and inclusion

## Notes

### Competing Interest Statement

The authors have declared no competing interest.

### Clinical Trial

NCT04789928

### Author Declarations

The trial was conducted in compliance with the principles of Good Clinical Practice and the Declaration of Helsinki for biomedical research involving human beings. It was approved by the French National Ethics Committee (Comite de Protection des Personnes, CPP Ile de France 11) and authorized by the Agence nationale de securite du medicament et des produits de sante. A written informed consent was obtained from all patients enrolled in the trial. The trial was registered at Clinical-Trials.gov (NCT04789928).

